# Multi-tract multi-symptom relationships in pediatric concussion

**DOI:** 10.1101/2021.04.01.21254814

**Authors:** Guido I. Guberman, Sonja Stojanovski, Eman Nishat, Alain Ptito, Danilo Bzdok, Anne Wheeler, Maxime Descoteaux

## Abstract

The heterogeneity of white matter damage and symptoms in concussion has been identified as a major obstacle to therapeutic innovation. In contrast, most diffusion MRI (dMRI) studies on concussion have traditionally relied on group-comparison approaches that average out heterogeneity. To leverage, rather than average out, concussion heterogeneity, we combined dMRI and multivariate statistics to characterize multi-tract multi-symptom relationships. Using cross-sectional data from 306 previously-concussed children aged 9-10 from the Adolescent Brain Cognitive Development Study, we built connectomes weighted by classical and emerging diffusion measures. These measures were combined into two informative indices, the first representing microstructural complexity, the second representing axonal density. We deployed pattern-learning algorithms to jointly decompose these connectivity features and 19 symptom measures. We found multivariate connectivity-symptom correspondences that were stronger than all single-tract single-symptom associations. Expression of multi-tract features was not driven by sociodemographic and injury-related variables. In a replication dataset, the expression of multi-tract features predicted psychiatric diagnoses after accounting for other psychopathology-related variables. These clinically-informative, cross-demographic multi-tract multi-symptom relationships recapitulated well-known findings from the concussion literature and revealed new insights about white matter structure/symptom relationships. These results may pave the way for the development of improved stratification strategies and the development of predictive biomarkers for personalized concussion management approaches.

## Introduction

Concussion afflicts approximately 600 per 100,000 individuals every year.^1^ Its incidence rate is rising in children and adolescents,^2^ and compared to adult populations, the impact of concussions on pediatric brains is understudied.^3^ Despite considerable funding devoted to clinical and basic research, no major advances in therapeutics have been achieved to date.^4^ A root cause of this stagnation appears to be a contradiction: while all concussions are treated equally in clinical trials and research studies, they are characterized by extensive heterogeneity in their pathophysiology, clinical presentation, symptom severity and duration.^4,5^ Concussion heterogeneity across patients has been identified as a major hurdle in advancing concussion care.^4,5^

Due to shearing forces transmitted during injury, the brain’s white matter is especially vulnerable to concussion.^6,7^ Decades of research have studied white matter structure in individuals who sustain concussions. However, most studies continue to assume consistent, one-to-one structure/symptom relationships and employ traditional group comparisons,^8,9^ averaging out the diffuse and likely more idiosyncratic patterns of brain structure abnormalities in favour of shared ones. Hence, the extant literature suggests that a large proportion of the clinical and research studies have not adequately accounted for clinical and neuropathological concussion heterogeneity.

To remedy this shortcoming, a growing number of studies aimed to parse the clinical heterogeneity in concussions by algorithmically partitioning patients into discrete subgroups based on symptoms.^10-12^ Other studies aim instead to account for heterogeneity in white matter structure alterations.^13-15^ Ware et al.^15^ built individualized maps of white matter abnormalities which revealed substantial inter-subject variability in traumatic axonal injury and minimal consistency of subject-level effects. Taylor et al.^14^ computed a multivariate summary measure of white matter structure across 22 major white matter bundles which achieved better classification accuracy of concussed patients from healthy controls compared to single tract measures. Hence, studies have attempted to address concussion heterogeneity in symptoms and in white matter structure. However, no prior studies have considered both sources of heterogeneity simultaneously.

White matter alterations due to concussion are diffuse and can elicit several symptoms that may interact with each other in complex ways.^4,5,16^ For instance, two individuals may suffer a concussion and develop sleep problems. The first may have damaged white matter tracts related to sleep/wakefulness control, whereas the second may have damaged tracts related to mood, causing depression-like symptoms, which include sleep problems. These two individuals will thus display a common symptom but will have overall different symptom profiles and different white matter damage profiles. Parsing concussion heterogeneity requires accounting for these dynamic, multi-tract multi-symptom relationships.

In the present study, we leveraged advanced diffusion MRI (dMRI) methods as well as a double-multivariate approach to parse concussion heterogeneity in white matter structure and symptoms simultaneously in a large sample of previously-concussed children. Multi-tract multi-symptom relationships captured more information than traditional univariate approaches. Expression of multi-tract connectivity features was not driven by sociodemographic strata and injury-related variables. Finally, after accounting for univariate variables found to be related to adverse psychiatric outcome in the discovery dataset (n=214), we found that expression of one multi-tract connectivity feature predicted adverse psychiatric outcome in a replication dataset (n=92).

## Results

### Sample

Out of 434 participants with a history of mTBI, 306 (127F/179M) had usable data (Figure 1). Table 1 outlines sociodemographic and injury-related factors, as well as handedness and sex. The majority had sustained an injury over 1 year prior to the study. Nuisance variables were well-balanced between participants in the discovery and the replication set.

**Table 1.** Table of sample characteristics. Note: Participants with “Unknown” Injury Mechanism and Total TBIs reported sustaining a TBI but no mechanism of injury was endorsed.

| **Demographic and injury data** | **Discovery set**<br>**(n=214)** | **Replication set**<br>**(n=92)** |
| --- | --- | --- |
| **Interview Age** |  |  |
| Mean (SD) | 9.57 (0.496) | 9.54 (0.501) |
| Median [Min, Max] | 10.0 [9.00, 10.00] | 10.0 [9.00, 10.0] |
| **Sex** |  |  |
| F | 88 (41.1%) | 39 (42.4%) |
| M | 126 (58.9%) | 53 (57.6%) |
| **Pubertal Stage** |  |  |
| Early | 41 (19.2%) | 18 (19.6%) |
| Mid | 58 (27.1%) | 19 (20.7%) |
| Prepubertal | 115 (53.7%) | 52 (56.5%) |
| Late | 0 (0%) | 3 (3.3%) |
| **Race/Ethnicity** |  |  |
| Asian | 2 (0.9%) | 2 (2.2%) |
| Hispanic | 27 (12.6%) | 18 (19.6%) |
| Multiple | 18 (8.4%) | 8 (8.7%) |
| Non-Hispanic Black | 14 (6.5%) | 11 (12.0%) |
| Non-Hispanic White | 151 (70.6%) | 52 (56.5%) |
| Other | 2 (0.9%) | 1 (1.1%) |
| **Combined Family Income** |  |  |
| <5K | 5 (2.3%) | 5 (5.4%) |
| \$5,000 - \$11,999 | 5 (2.3%) | 1 (1.1%) |
| \$12,000-\$15,999 | 3 (1.4%) | 2 (2.2%) |
| \$16,000-\$24,999 | 5 (2.3%) | 3 (3.3%) |
| \$25,000-\$34,999 | 12 (5.6%) | 4 (4.3%) |
| \$35,000-\$49,999 | 12 (5.6%) | 5 (5.4%) |
| \$50,000-\$74,999 | 34 (15.9%) | 16 (17.4%) |
| \$75,000-\$99,999 | 31 (14.5%) | 13 (14.1%) |
| \$100,000-\$199,000 | 76 (35.5%) | 27 (29.3%) |
| >\$200,000 | 31 (14.5%) | 16 (17.4%) |
| **Handedness** |  |  |
| LH | 10 (4.7%) | 10 (10.9%) |
| RH | 175 (81.8%) | 69 (75%) |
| Mixed | 29 (13.6%) | 13 (14.1%) |
| **Injury Mechanism** |  |  |
| Fall/hit by object | 135 (63.1%) | 48 (52.2%) |
| Fight/shaken | 2 (0.9%) | 3 (3.3%) |
| Motor vehicle collision | 14 (6.5%) | 3 (3.3%) |
| Multiple | 10 (4.7%) | 5 (5.4%) |
| Unknown | 53 (24.8%) | 33 (35.9%) |
| **Time Since Injury** |  |  |
| Mean (SD) | 3.22 (2.79) | 3.23 (2.60) |
| Median [Min, Max] | 2.00 [0.00, 11.0] | 2.50 [0.00, 9.00] |
| **Total TBIs** |  |  |
| Unknown | 53 (24.8%) | 33 (35.9%) |
| 1 | 151 (70.6%) | 54 (58.7%) |
| 2 | 9 (4.2%) | 5 (5.4%) |
| 3 | 1 (0.5%) | 0 (0%) |

**Figure 1.**
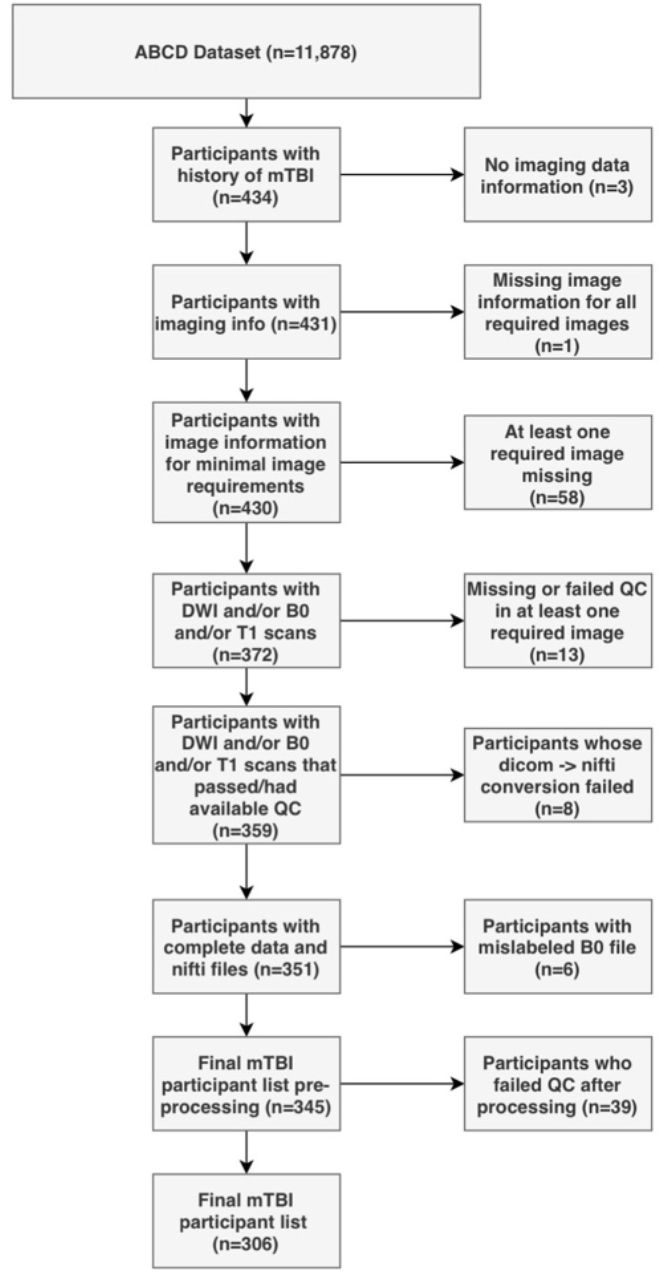
Flowchart describing the participant selection procedure.

### Combined measures of white matter tract microstructure

The PCA yielded two biologically-interpretable components that together explained 97% of the variance in dMRI measures (Figure S2). The first appeared to reflect an index of microstructural complexity, whereas the second more closely reflected axonal density. Because we retained two PCs, we performed two PLSc analyses.

### Multi-tract multi-symptom relationships

Each PLSc analysis yielded 19 latent modes of covariance (termed here “multi-tract multi-symptom relationships”), each consisting of a pair of multi-tract connectivity and multi-symptom features. Based on permutation testing, 18 multi-tract multi-symptom pairs were retained from the microstructural complexity PLSc, and 14 from the axonal density PLSc. For brevity, only a few informative pairs are presented herein, but an illustration of all significant pairs for all thresholds can be found in supplementary material (Figures S3-S6).

For microstructural complexity, similarly to axonal density, the first multi-tract multi-symptom pair broadly represented all symptoms (Figure 2A), capturing general problems. Interestingly, this pair implicated several callosal tracts (Figure 3). The third multi-tract multi-symptom pair obtained from the axonal density PLSc represented mostly sleep and cognitive problems, whereas the fourth pair represented more strongly mood, sleep, and somatic problems (Figure 2B and C).

**Figure 2.**
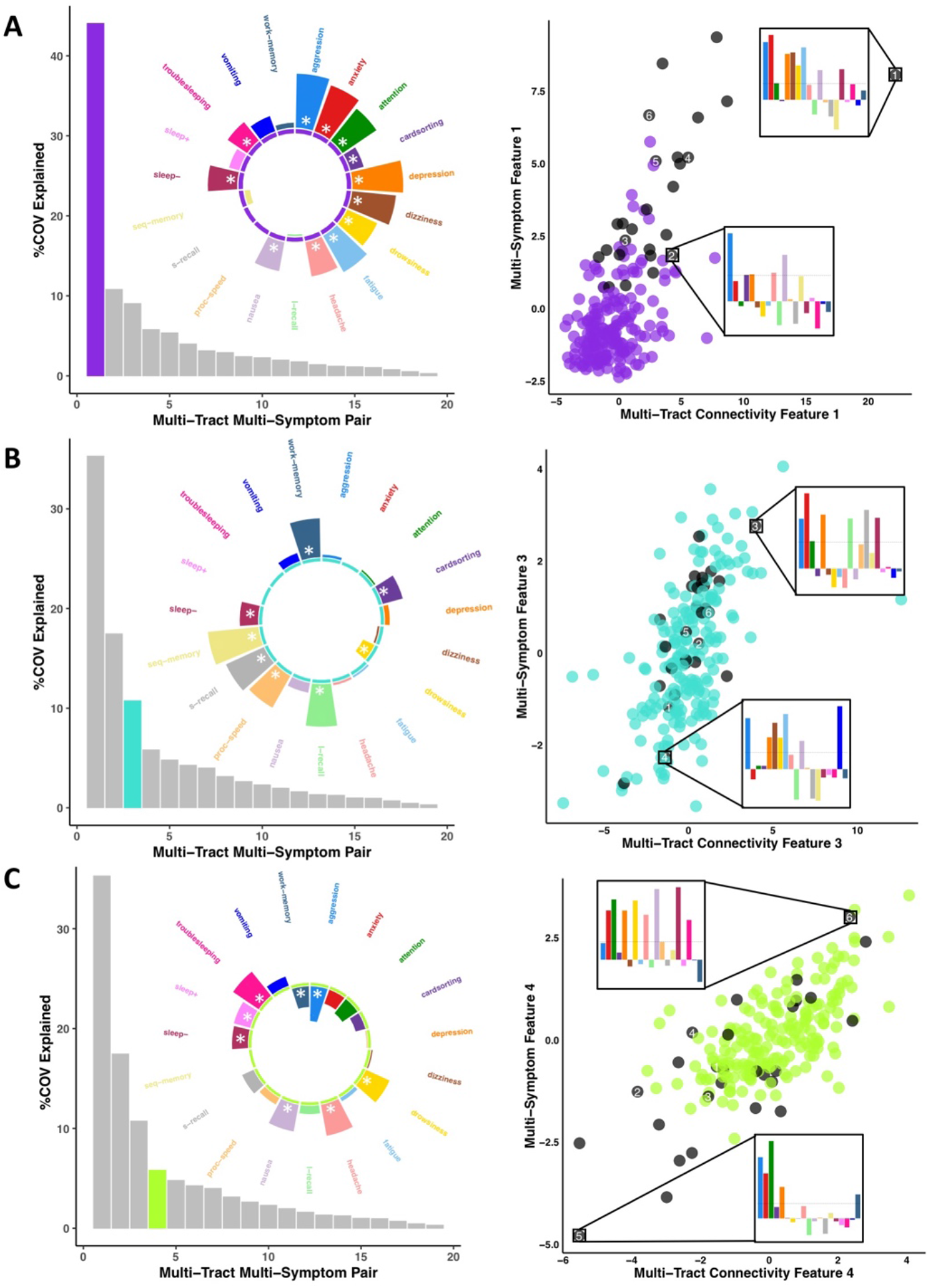
Illustration of 3 multi-tract multi-symptom pairs obtained from the microstructural complexity PLSc (A), and from the axonal density PLSc (B and C). Left: Polar plots displaying the weights of all 19 symptom measures for each multi-symptom feature. Each circle is color-coded to match the corresponding scatter plot. Bars pointing away from the center illustrate positive weights, bars pointing towards the center represent negative weights. White stars illustrate symptoms that significantly contributed to the pair. Bar graphs underneath the polar plots illustrate the % covariance explained by each pair, with the currently-shown pair highlighted. Right: Scatter plots showing the expression of multi-tract features (x-axis) and multi-symptom features (y-axis). In each plot, the same 6 participants are labelled (1 through 6), with each plot showing the scaled symptom *measures* (i.e.: not the expression of multi-symptom features) for two participants, one expressing low levels of a pair, the other expressing high levels. For each illustrated participant, positive bars illustrate symptoms that are higher than the sample average, negative bars represent symptoms that are lower. Participants with clinical levels of CBCL Depression, Attention Problems, Anxiety Disorder, or Aggression scores are illustrated in black. A. Illustration of multi-tract multi-symptom pair 1 obtained from the microstructural complexity PLSc. B. Illustration of multi-tract multi-symptom pair 3 obtained from the axonal density PLSc. C. Illustration of multi-tract multi-symptom pair 4 obtained from the axonal density PLSc.

**Figure 3.**
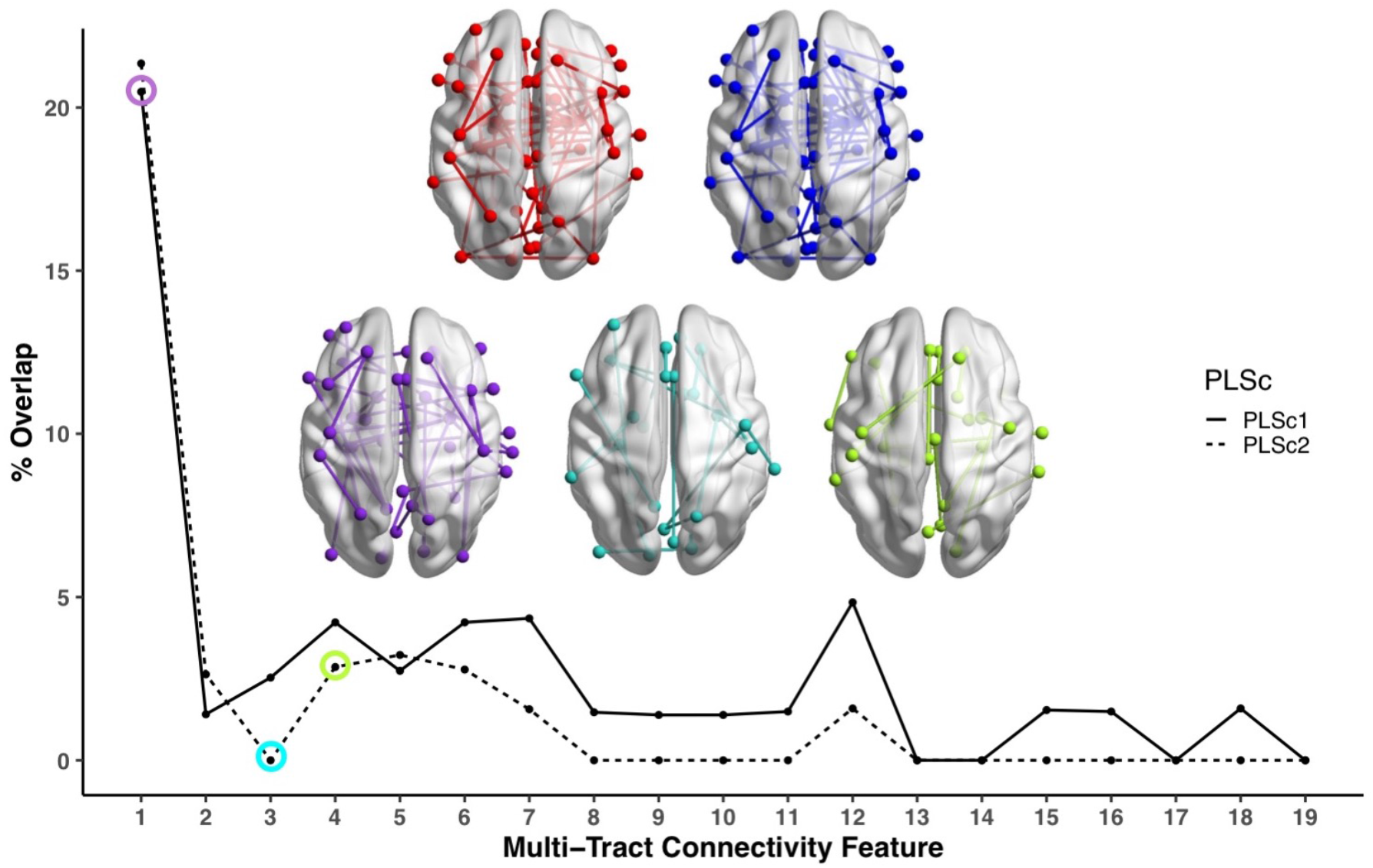
Line plot showing the percent overlap between univariate analyses and each multi-tract connectivity feature. Highest overlap occurred for the first multi-tract connectivity feature from both PLSc analyses. Brain renderings shown above graph illustrate which connections were found to be significant for univariate comparisons of microstructural complexity (red), univariate comparisons of axonal density (blue), multi-tract connectivity feature 1 from the microstructural complexity PLSc (violet), multi-tract connectivity feature 3 from the axonal density PLSc (turquoise), and multi-tract connectivity feature 4 from the axonal density PLSc (green). These brain renderings illustrate how many callosal connections are present for univariate comparisons and for multi-tract connectivity feature 1, but less so for multi-tract connectivity features 3 and 4. The percent overlap score for each of the three illustrated multi-tract connectivity features are identified in the line plot with a circle of the corresponding color. Brain renderings were visualized with the BrainNet Viewer.^17^

### Multivariate vs univariate approaches

Correlations between multi-tract and multi-symptom features were higher than all univariate correlations between the microstructural complexity and axonal density scores of every retained tract and every symptom (“single-tract single-symptom relationships”).

To further explore this comparison between univariate and multivariate approaches, we identified 28 individuals with scores above 70 in CBCL Depression, Attention Problems, Anxiety Disorder, or Aggression scales, considered to be the clinical range.^9^ Figure 3 (scatter plots) illustrates the expression of three multi-tract multi-symptom pairs. In clinical trials and research studies, these 28 individuals would be grouped together. However, these individuals (displayed in black) were differentiable by their expression of multi-tract and multi-symptom features (Figure 2).

We compared microstructural complexity and axonal density scores across all 200 connections between individuals with and without clinical-level CBCL scores, and calculated the percent overlap between each multi-tract connectivity feature and the set of tracts found to be significant in univariate comparisons. The percent overlap scores are presented in Figure 3. Notably, the highest overlap occurred with multi-tract connectivity feature 1 (22%) from both PLSc analyses. Univariate psychopathology-related features (Figure 3 red and blue brain renderings) and multi-tract connectivity feature 1 (Figure 3 violet brain rendering) implicated several callosal tracks, whereas fewer callosal tracks were implicated in multi-tract connectivity features 3 and 4 (Figure 3 turquoise and green brain renderings).

### Relationship with sociodemographic and injury-related factors

The expression of multi-tract connectivity features was equivalent across sociodemographic strata defined by sex, total combined household income, and race/ethnicity (Figure S1). Out of 32 retained multi-tract multi-symptom pairs, time since the latest injury was only significantly correlated to the expression of two multi-tract connectivity features (Table S1) and no multi-symptom features. Only the expression of one multi-tract connectivity feature (feature 2 from the microstructural complexity PLSc) was significantly different between groups defined by injury cause (Figure S7). Only the expression of two multi-tract connectivity features, feature 17 from the microstructural complexity PLSc (Figure S8), and feature 8 from the axonal density PLSc (Figure S9) significantly differed between groups defined by the total number of TBIs.

### Prediction of clinical outcome

118 children (55%) in the discovery set and 51 (55%) in the replication set had an adverse psychiatric outcome. Using separate multivariable logistic regressions, we found expression of four multi-tract connectivity features from the microstructural complexity PLSc, one behavioural measure (CBCL Attention Problems), and no single tract features that were significantly related to adverse psychiatric outcome. No other variables, including no multi-tract connectivity features obtained from the axonal density PLSc, were related to adverse psychiatric outcome. We computed microstructural complexity scores for the replication set, selected the same 200 connections that had been retained in the discovery dataset, and computed the expression of the multi-tract connectivity features that had been found to be significantly related to adverse psychiatric outcome in the discovery set. We incorporated these multi-tract connectivity features with the other significant variables, using the replication data, in a single multivariable logistic regression. In this model, we also added time since the latest injury. We found that a one-unit increase in the expression of multi-tract connectivity feature 4 significantly increased the odds of an adverse psychiatric outcome in the replication set 2.50 times (*p*=0.01) after controlling for univariate, psychopathology-related behavioural features and time since the latest injury. This multi-tract connectivity feature was paired with a multi-symptom feature that implicated sleep and somatic problems (Figure S10).

### Sensitivity analyses

Results using different thresholds (t=85%, 95%, 100%) for the connectomes are outlined in supplementary material. All results were consistent, including PCs, the weights of the multi-symptom features, the strength of multivariate vs univariate relationships, the trends for the % overlap scores, and the significance of adverse psychiatric outcome predictions using multi-tract connectivity feature expression. Results were less consistent with the t=100% threshold, which demonstrated instead little to no significant multi-tract multi-symptom pairs based on permutation testing. Correlations between expression of multi-tract connectivity features at different thresholds are illustrated in supplementary material. Notably, for t=85%, 90%, and 95%, correlations between the expression of the corresponding multi-tract connectivity features (e.g.: multi-tract feature 1 from t=85%,multi-tract feature 1 from t=95%) all have high correlations, indicating how similar these features are, whereas these correlations were lower when comparing against t=100% (Figure S11).

## Discussion

In the present study we leveraged novel dMRI methods and a double-multivariate approach to parse heterogeneity in white matter structure and symptoms in a large sample of previously-concussed children. By applying PLSc on biologically-interpretable measures of dMRI obtained from PCA, we found cross-demographic multi-tract multi-symptom relationships. These multi-tract multi-symptom pairs captured more information than traditional approaches and predicted meaningful clinical outcomes in unseen data. Additionally, these results recapitulated well-known findings from the concussion literature and revealed new insights about white matter structure/symptom relationships.

Concussion heterogeneity has been identified as a major obstacle^4,5^ in response to decades of failed attempts to translate basic science findings into successful clinical trials and novel therapies. Heterogeneity in symptoms, impact of injury on brain structure and function, and pre-injury factors pose a particular problem for most concussion neuroimaging studies which have traditionally employed univariate comparisons between concussed and healthy or orthopedic injury control groups, or between patients with and without persistent symptoms.^8,9^ These sources of variability are believed to be problematic because they decrease the statistical power needed for group comparisons and multivariable models to detect the often-subtle effects of concussions.^18^ To overcome this challenge, landmark initiatives such as the IMPACT,^18^ InTBIR,^19^ CENTER TBI,^20^ and TRACK TBI^21^ aim to standardize and pool multi-center data collected across sociodemographic strata, to identify and statistically correct for pre-injury factors known to impact brain structure, and develop diagnostic and prognostic tools leveraging multimodal data and increasingly sophisticated machine-learning approaches.

In this study, we posited that concussion heterogeneity is also problematic because by pooling across patients, idiosyncratic patterns of connectivity that may be more symptom-specific are sacrificed in favour of shared ones. By assuming that symptoms map cleanly and consistently onto shared connectivity abnormalities in a one-to-one fashion, erroneous inferences could be made about relationships between group-level patterns of connectivity differences and specific symptoms. Our results are consistent with this idea: univariate comparisons between a group displaying clinical-level psychopathology and the rest of the sample identified connectivity features that mostly overlapped with the first multi-tract multi-symptom pair obtained from both PLSc analyses. These pairs, which accounted for the most covariance, reflected general problems and not specifically psychopathology. Both these pairs and the univariate “psychopathology-related” connections implicated several callosal tracts. These results suggest that univariate comparisons, even when performed in such a way as to identify a symptom-specific set of connectivity features, identified only the most consistent group-level connectivity differences at the expense of more symptom-specific idiosyncratic ones. These findings are consistent with most prior concussion literature, which strongly implicate the corpus callosum,^9^ but extend this literature by suggesting that that the central importance of the corpus callosum may have resulted from pooling across concussed subjects.

Rather than a clean and consistent set of single-tract single-symptom relationships, this study suggests concussions may be best conceptualized as a multiplicity of multi-tract multi-symptom combinations. Multi-tract relationships may be driven by the metabolic demands imposed by the network structure of the brain, which is known to predict the course of several brain diseases,^22^ by biomechanical constraints imposed by the skull and other structures exposing certain areas to more shearing strain,^23^ or by both factors simultaneously.^24^ These possibilities need to be tested further.

After accounting for behavioural and single-tract predictors of adverse psychiatric outcome, the expression of one multi-tract connectivity feature significantly predicted adverse psychiatric outcome in a holdout dataset. This multi-tract feature was paired with a multi-symptom feature that implicated mostly sleep and somatic problems. Sleep problems are known to be linked to psychiatric disorders, both as symptoms, and risk factors.^25^ Longitudinal studies are needed to elucidate whether these multi-tract features reflect abnormalities that increase the risk of later adverse psychiatric outcomes, or instead reflect abnormalities associated with existing psychiatric diagnoses. Nonetheless, the prediction of a meaningful clinical outcome on unseen data is particularly promising.

The present findings must be contrasted to the nascent literature addressing concussion heterogeneity. A few recent studies have parsed heterogeneity in concussion symptoms, all using clustering analyses.^10-12^ The reported subgroups differed from those found in the present study. Differences between symptom profiles arose because our multi-symptom features are associated with brain structure and not driven by variability in symptoms alone. Further, we did not group patients into discrete subgroups. Although whether patients cleanly fit into discrete subtypes has not been explicitly tested in our study, the expression of multi-tract and multi-symptom features do not suggest any obvious clusters (e.g.: Figure 3 scatter plots). The possibility of concussion subtypes needs to be explored further. Other prior studies have attempted to address heterogeneity in white matter structure in concussions.^13-15^ Using different approaches, these studies generated point summaries that accounted for the high-dimensional variability of white matter structure to better distinguish patients from controls. Our approach offers an important advantage. Rather than collapsing the rich and highly variable information provided by white matter structure, our approach attempts to find symptom-specific patterns of white matter structure, which has potential to lead to new treatment targets.

The present results should be considered in light of methodological limitations. Data on mTBI occurrence was collected retrospectively. Participants did not have baseline data, and additionally had highly variable times since injury. Most individuals with concussions recover from their injury^26^ which should have led to a concussed group where most participants were similar to healthy controls. Interestingly, our PCA yielded combinations of diffusion measures that differed from those of two prior studies that have used this approach.^27,28^ These prior studies used samples of typically-developing children without neurological insults. To the extent that the PCs reported in these prior studies reflect healthy neurotypical brains, our PCs suggest that our concussed sample was not as similar in white matter structure to healthy controls as expected. However, variable time since injury made the interpretation of patterns of microstructure difficult. Further, due to the cross-sectional nature of this data, the difference between symptoms and pre-existing characteristics are difficult to discern, especially since some behavioural measures often believed to be symptoms of concussion, such as attention problems, can also be risk factors for injury.^29^ Lastly, heterogeneity has several forms, including in symptoms, duration, severity, neuropathology,^7^ lesion location,^15^ sociodemographics,^18^ genetics,^30^ behaviour,^29^ pre-injury comorbidities,^31^ and environmental differences, including access to and quality of care.^32^ These factors have been theorized to interact in complex ways.^4^ This study only addressed a minority of these complex relationships, further studies integrating more variable sets are needed to address these other drivers of heterogeneity.

Conversely, this study leveraged some of the most recent and important advances in dMRI to address the major limitations of conventional approaches. We used high quality multi-shell dMRI data,^33^ as well as modelling approaches, tractography techniques, and microstructural measures robust to crossing fibers, partial volume effects, and connectivity biases.^27,34-38^ We used PCA to combine dMRI measures into meaningful indices of white matter structure. Lastly, we used gold-standard measures of psychiatric illness to predict clinical outcomes. Future iterations of this work will need better control of time since injury, longitudinal follow-up, broader assessment of clinical outcomes, and further development of predictive models.

In conclusion, leveraging advanced dMRI and a pattern-learning algorithm to parse concussion heterogeneity, we have found clinically-meaningful, cross-demographic multi-tract multi-symptom relationships. As the field moves towards large-scale studies which aim to statistically control for sociodemographic sources of heterogeneity to detect a putative consistent white matter signature of concussion across patients, the insights gained from this study should be taken into consideration: informative, clinically-meaningful, symptom-specific patterns of connectivity differences are lost when pooling across concussed patients. This insight is an important step towards improving stratification strategies for clinical trials, identifying novel treatment targets, and developing predictive biomarkers for personalized concussion management approaches.

## Materials and Methods

### Participants

Participants in this study were obtained from the world’s largest child development study of its kind – the ongoing longitudinal Adolescent Brain Cognitive Development Study (ABCD Study; https://abcdstudy.org/), data release 2.0 (https://data-archive.nimh.nih.gov/abcd). The ABCD Study acquired data from 11,874 children aged 9 to 10 years (mean age = 9.49 years) from across the United States (48% girls; 57% Caucasian, 15% African American, 20% Hispanic, 8% other).^39^ Additional information about the ABCD Study can be found in Garavan et al.^40^

### History of concussion

Parents completed a modified version of the Ohio State University TBI Identification Method (OSU-TBI-ID) ^41^. We included participants who reported a head injury without loss of consciousness but with memory loss and/or a head injury with loss of consciousness for less than 30 minutes (n=434). Due to missing or incomplete data, corrupted files, data conversion errors, and images rated by the ABCD Study team as being of poor quality, the final sample of participants with usable data was 345. After processing, images were visually inspected by two trained independent raters (G.I.G., S.S.). Images that were deemed of low quality after processing by both raters were removed (n=39), leading to a final sample of 306 participants. We randomly divided the sample into a discovery dataset (70%, n=214) and a replication dataset (20%, n=92). Figure 1 summarizes the subject selection procedure.

### Symptom-oriented measures

To probe various aspects of concussion symptomatology, we used items collected from assessments available in the ABCD dataset. These items, as well as the concussion symptom they are meant to probe are outlined in Table 2.

**Table 2.**
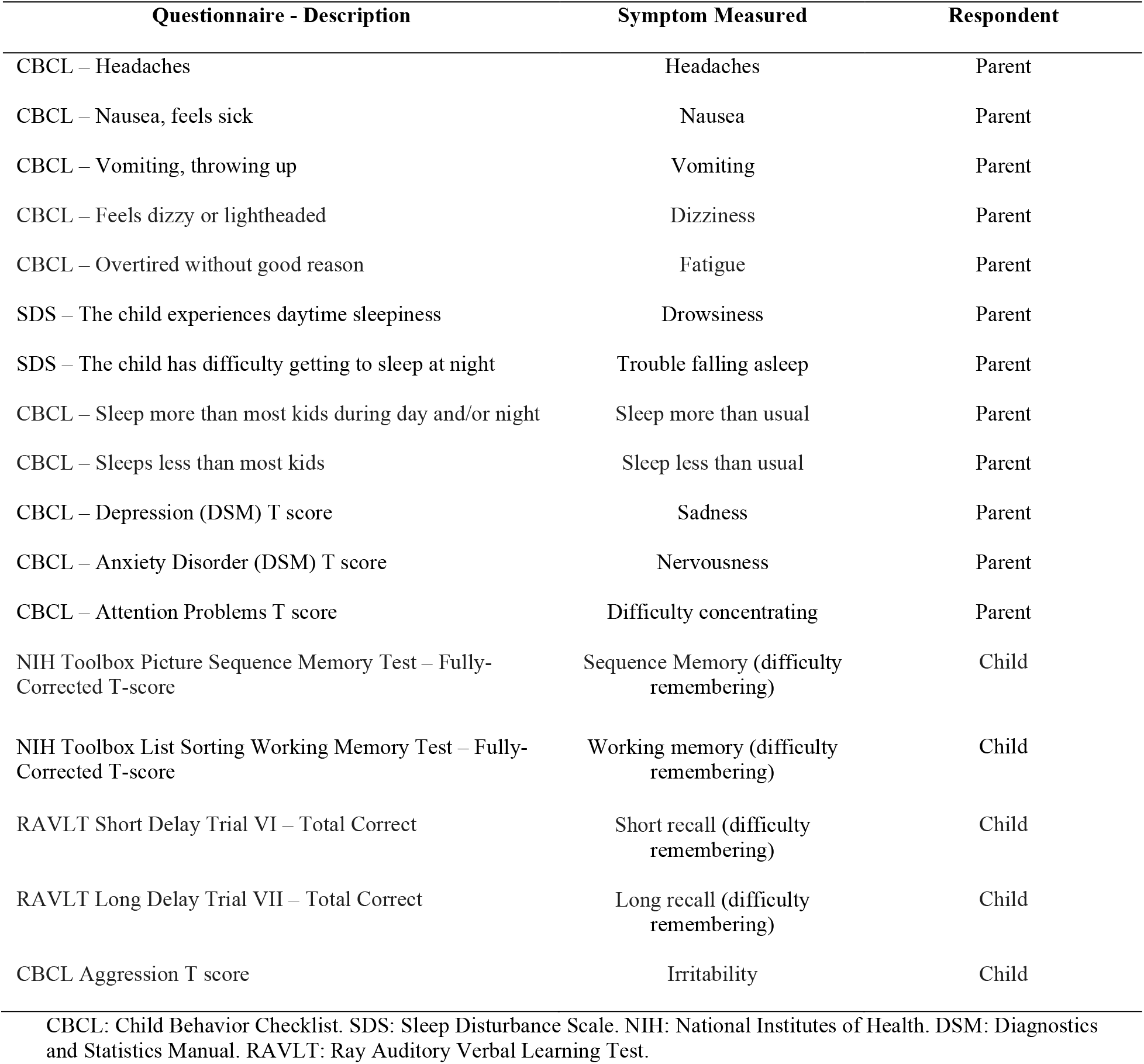
Table outlining all behavioural measures used in analyses, along with the corresponding symptom they reflect.

| Questionnaire - Description | Symptom Measured | Respondent |
| --- | --- | --- |
| CBCL – Headaches | Headaches | Parent |
| CBCL – Nausea, feels sick | Nausea | Parent |
| CBCL – Vomiting, throwing up | Vomiting | Parent |
| CBCL – Feels dizzy or lightheaded | Dizziness | Parent |
| CBCL – Overtired without good reason | Fatigue | Parent |
| SDS – The child experiences daytime sleepiness | Drowsiness | Parent |
| SDS – The child has difficulty getting to sleep at night | Trouble falling asleep | Parent |
| CBCL – Sleep more than most kids during day and/or night | Sleep more than usual | Parent |
| CBCL – Sleeps less than most kids | Sleep less than usual | Parent |
| CBCL – Depression (DSM) T score | Sadness | Parent |
| CBCL – Anxiety Disorder (DSM) T score | Nervousness | Parent |
| CBCL – Attention Problems T score | Difficulty concentrating | Parent |
| NIH Toolbox Picture Sequence Memory Test – Fully-Corrected T-score | Sequence Memory (difficulty remembering) | Child |
| NIH Toolbox List Sorting Working Memory Test – Fully-Corrected T-score | Working memory (difficulty remembering) | Child |
| RAVLT Short Delay Trial VI – Total Correct | Short recall (difficulty remembering) | Child |
| RAVLT Long Delay Trial VII – Total Correct | Long recall (difficulty remembering) | Child |
| CBCL Aggression T score | Irritability | Child |

CBCL: Child Behavior Checklist. SDS: Sleep Disturbance Scale. NIH: National Institutes of Health. DSM: Diagnostics and Statistics Manual. RAVLT: Ray Auditory Verbal Learning Test.

### MRI Acquisition

MRI scans were acquired across 21 sites, with data coming from 28 different scanners. Details about the acquisition protocols and image specifications are outlined in Casey et al 2018.^42^ Multi-shell dMRI scans had 96 diffusion-weighted directions, with 6 directions of b=500 s/mm^2^, 15 directions of b=1000s/mm^2^, 15 directions of b=2000s/mm^2^, and 60 directions of b=3000s/mm^2^. The b=2000 shell was excluded from the data processing. In addition, scans had 6 or 7 b=0s/mm^2^ images, depending on scanner type. Lastly, a reverse b0 image was included for each participant.

### Processing

We used Tractoflow^43^ to process dMRI and T1-weighted scans. Tractoflow is a novel diffusion MRI processing pipeline, incorporating state-of-the-art functions from FSL, Dipy, and MRtrix into NextFlow. The processing steps are summarized in Theaud et al.^43^ Important deviations from the default parameters utilized by Tractoflow are as follows: 1. We used gray-white matter interface seeding, as this method accounts for the length bias introduced by white-matter seeding;^36^ 2. We used 24 seeds-per-voxel with the objective of obtaining approximately 2 million streamlines across the entire brain. We used the b=0, 500, and 1000 shells to perform tensor fitting, and the b=0 and 3000 shells to perform Constrained Spherical Deconvolution (CSD).^35,38^ We obtained group-average fiber-response functions from voxels with high (>0.70) fractional anisotropy (FA). Lastly, we created tractograms using a probabilistic particle-filtering tractography algorithm.^36^

### Connectivity matrices

The post-processing workflow is illustrated in Figure 4. To construct connectivity matrices, we used Freesurfer on McGill’s CBrain platform^44^ to fit the Desikan-Killiani Tourvile (DKT)^45^ and *aseg* atlases onto the processed T1-images that had been transformed to DWI space during processing (Figure 4A). We applied these parcellations and extracted diffusion measures using *connectoflow* (https://github.com/scilus/connectoflow). We removed redundant and irrelevant labels from the fitted atlas (a list of retained labels is supplied in supplementary material), yielding a final atlas with 76 labels. We then thresholded matrices such that a connection was only retained if it was found to be successfully reconstructed (defined as the presence of at least one streamline) across 90% of participants.^46^ Results using different thresholds are presented in supplementary material. We then weighted thresholded connectomes by FA, mean, radial, and axial diffusivities (MD, RD, AD respectively), apparent fiber density along fixels (AFDf), and number of fiber orientations (NuFO) (Figure 4B). The first four measures are derived from the tensor model, whereas the latter two are based on fiber orientation distribution functions (fODFs) obtained from CSD.^34,37^ Simulation studies have shown that AFD is more specifically related to axonal density, and by computing it along “fixels” (fiber elements), axonal density specific to particular fiber populations can be studied independently of crossing fibers.^37^ Although individual diffusion measures are related to different aspects of neuropathology, together they provide more information than when considered separately.^46^ A recent framework based on principal component analysis (PCA) has been proposed to combine diffusion measures into biologically-interpretable indices of white matter structure.^27^ We therefore performed PCA on the concatenated set of standardized measures across subjects and connections, generating connectivity matrices weighted by principal component (PC) scores (Figure 4C).

**Figure 4.**
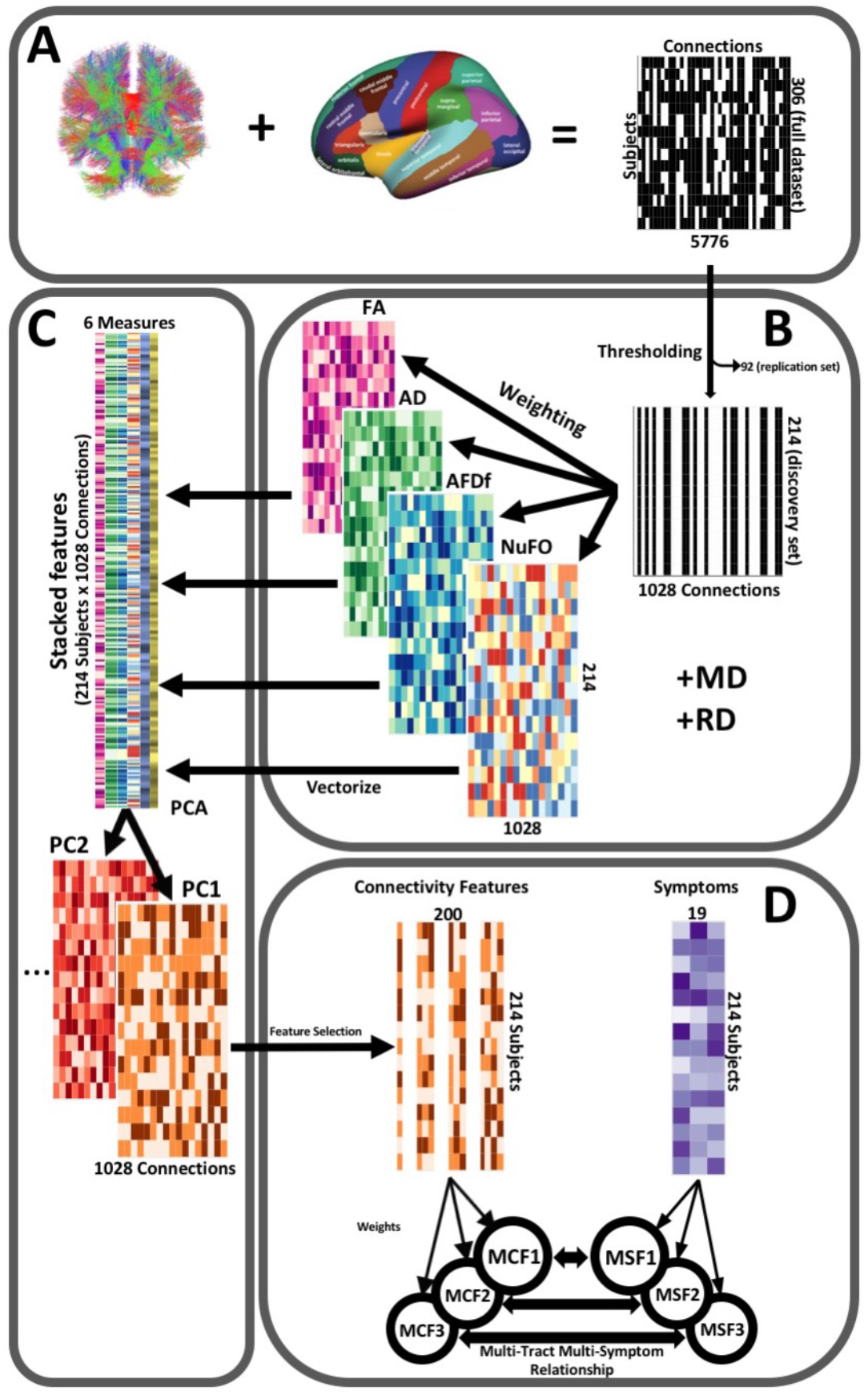
Illustration of the study’s post-processing pipeline. A. We applied the DKT parcellation onto each tractogram, thus building a binary connectivity matrix that displayed for all 306 subjects in the full dataset (rows), whether (black) or not (white) a streamline existed between each pair of labels (columns). B. We split the dataset into a discovery set (n=214) and a replication set (n=92). On the discovery set, we thresholded connectomes, only keeping connections that existed across 90% of participants (a threshold of 100% is illustrated here for simplicity). We then constructed connectomes of 6 scalar diffusion measures (Fractional Anisotropy (FA), Axial Diffusivity (AD), Mean Diffusivity (MD), Radial Diffusivity (RD), Apparent Fiber Density along fixels (AFDf), and Number of Fiber Orientations (NuFO)), by computing the average measure across each connection. C. We stacked all columns from each connectivity matrix, creating vectors of every pair of subject and connection, and then joined together these vectors. We then performed principal component analysis (PCA) on these matrices. Principal component (PC) scores were calculated for each subject/connection combination, thus reconstructing connectomes weighted by PC scores. D. From each these new connectomes, we selected 200 connections based on Pearson correlations with symptom-oriented measures. We then performed partial least squares correlation on each of these PC-weighted features and symptom measures, which allowed us to obtain pairs of multi-tract connectivity features (“MCF”) and multi-symptom features (“MSF”). Each multivariate feature is composed of linear combinations (weighted sums, illustrated by the black arrows called “weights”) of variables from its corresponding feature set.

### Additional data transformations

We imputed missing connectivity (prior to the PCA), symptom, and nuisance data (sex, pubertal stage, handedness, scanner) by randomly selecting non-missing data from other participants in the same dataset. We reverse-coded cognitive scores, such that increasing scores in all symptom data reflected more problems. From connectivity and symptom data, we regressed out the following nuisance variables: sex, pubertal stage, scanner (only for connectivity data), and handedness.

### Pattern-learning pipeline

#### Feature selection

To reduce the number of connectivity features included in the partial least squares correlation (PLSc) analysis, we selected the 200 connectivity features most correlated (based on Pearson correlations) with any symptom score. This solution is becoming increasingly adopted for high-dimensional variable sets (Figure 4D).^47,48^

#### PLSc

We performed PLSc analyses using the *tepPLS* function from the *texposition* package.^49^ PLSc involves singular value decomposition on the covariance matrix between connectivity and symptom features, creating pairs of multi-tract and multi-symptom features called multi-tract multi-symptom relationships. Each multi-tract multi-symptom relationship encapsulates a linear combination of connectivity features (“multi-tract features”), a linear combination of symptom scores (“multi-symptom features”), and an eigenvalue (reflective of the amount of explained covariance between connectivity and symptom features). Each multi-tract multi-symptom relationship is constructed so as to explain a successively smaller portion of the covariance between symptoms and connectivity features. We constructed the largest number of possible multi-tract multi-symptom relationships, given the dimensionality of the behavioral variable set (k=19) (Figure 4D).

### Selection and interpretation of multi-tract multi-symptom pairs

To select the multi-tract multi-symptom pairs to retain for interpretation, we performed permutation testing (2000 iterations). This procedure randomly shuffles row labels for the connectivity features, without replacement, repeats the PLSc and computes eigenvalues at every permutation. We calculated *p*-values as the proportion of permutations that yielded eigenvalues that exceeded the original amount.

To interpret symptom and connectivity weights of significant (*p*<0.05) multi-tract multi-symptom pairs, we performed bootstrap analyses (2000 iterations), using the *BOOT4PLSC* command from the *texposition* package. At each iteration, labels for data were drawn with replacement, the entire PLSc was repeated and the weights for all pairs were obtained. This process yields a sampling distribution of weights for each connectivity and symptom feature.^50^ The ratio of the original weights to the standard error of each feature’s bootstrap distribution can be interpreted as a *z*-score, which yielded so-called ‘bootstrap ratios’. We used a value of 1.96 to determine which variables significantly contributed to each particular significant pair.

### Comparison of multivariate against univariate approaches

To compare information captured by the PLSc and univariate approaches, we first divided participants based on whether they had T-scores above 70, a well-recognized clinical threshold,^51^ in the Child Behavior Checklist (CBCL) Depression, Attention Problems, Anxiety Disorder, or Aggression scales.^51^ Using a threshold of *p*<0.05, we computed univariate comparisons of connectivity (PC scores) between individuals with and those without clinical-level psychopathology, thus identifying psychopathology-related univariate connectivity features.

We were interested in comparing how many of these features were also found to significantly contribute to each multi-tract connectivity feature. To do so, we computed a measure of percent overlap as follows:

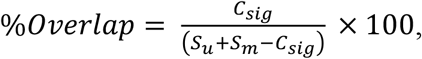

where C_sig_ refers to the number of connections flagged as significant in both approaches, S_u_ to the number of connections flagged as significant in the univariate approach, and S_m_ to the number of connections flagged as significant in the multivariate approach. This measure can account for the apparent high overlap that can arise when S_u_ and S_m_ are not equivalent in size.

### Relation to TBI-related and sociodemographic factors

We addressed whether expression of multi-tract connectivity features was related to injury-specific and sociodemographic factors. Injury-related variables included: the time between the last-documented injury and testing, the cause of injury, and the total number of documented mTBIs. Sociodemographic variables included: sex, total combined family income in the last 12 months, and race/ethnicity. We used the following categories for race/ethnicity: “Asian” (Asian Indian, Chinese, Filipino, Japanese, Korean, Vietnamese, Other Asian), AIAN (“American Indian”/Native American, Alaska Native), NHPI (Native Hawaiian, Guamanian, Samoan, Other Pacific Islander), Non-Hispanic White, Non-Hispanic Black, Hispanic, Other, and Multiple.^52^ To illustrate the influence of these sociodemographic factors we created scatter plots illustrating expression of connectivity latent factors color-coded by sociodemographic factors (Figure S1).

### Prediction of clinical outcome

We used the Kiddie-Schedule for Affective and Psychiatric Disorders in School Age Children (KSADS), a gold-standard tool, to assess the presence of pediatric psychiatric disorders.^53^ We used the presence of any current psychiatric diagnosis as indicating an adverse psychiatric outcome (55% of discovery set, 55% of replication set). We created separate multivariable logistic regressions to predict adverse psychiatric outcomes using 1) expression of multi-tract connectivity features, 2) psychopathology measures from the CBCL, or 3) psychopathology-related univariate connectivity features. From each set of models, we retained the variables that were significantly related to adverse psychiatric outcome. We then tested whether expression of multi-tract connectivity features could predict adverse psychiatric outcome in the replication dataset.

To do so, we first projected connectivity data from the replication dataset onto the principal components retained in the discovery dataset. Next, we selected the same 200 connections that had been retained based on univariate feature selection, and projected these connections onto the multi-tract connectivity spaces obtained from the discovery dataset. Finally, we performed a logistic regression model incorporating all the predictors found to be significant in the discovery dataset as well as the time since injury, applied on data from the replication dataset.

The objective of these models was to assess whether expression of multi-tract connectivity features could predict adverse psychiatric outcomes in the replication dataset after accounting for other psychopathology-related variables (CBCL behavioural measures, univariate connectivity measures) as well as time since injury.

### Data availability

Data from the ABCD Study is publicly available, and all scripts used in this study are openly-available as well (see references).

## Data Availability

Data for this manuscript was obtained from the Adolescent Brain Cognitive Development (ABCD) study. This dataset is openly available.

## Acknowledgments

The authors would like to thank François Rheault, Derek Beaton, Manon Edde, Guillaume Theaud, and Arnaud Boré for comments and other contributions that were helpful in this project.

## Supplementary Material

**Figure S1.**
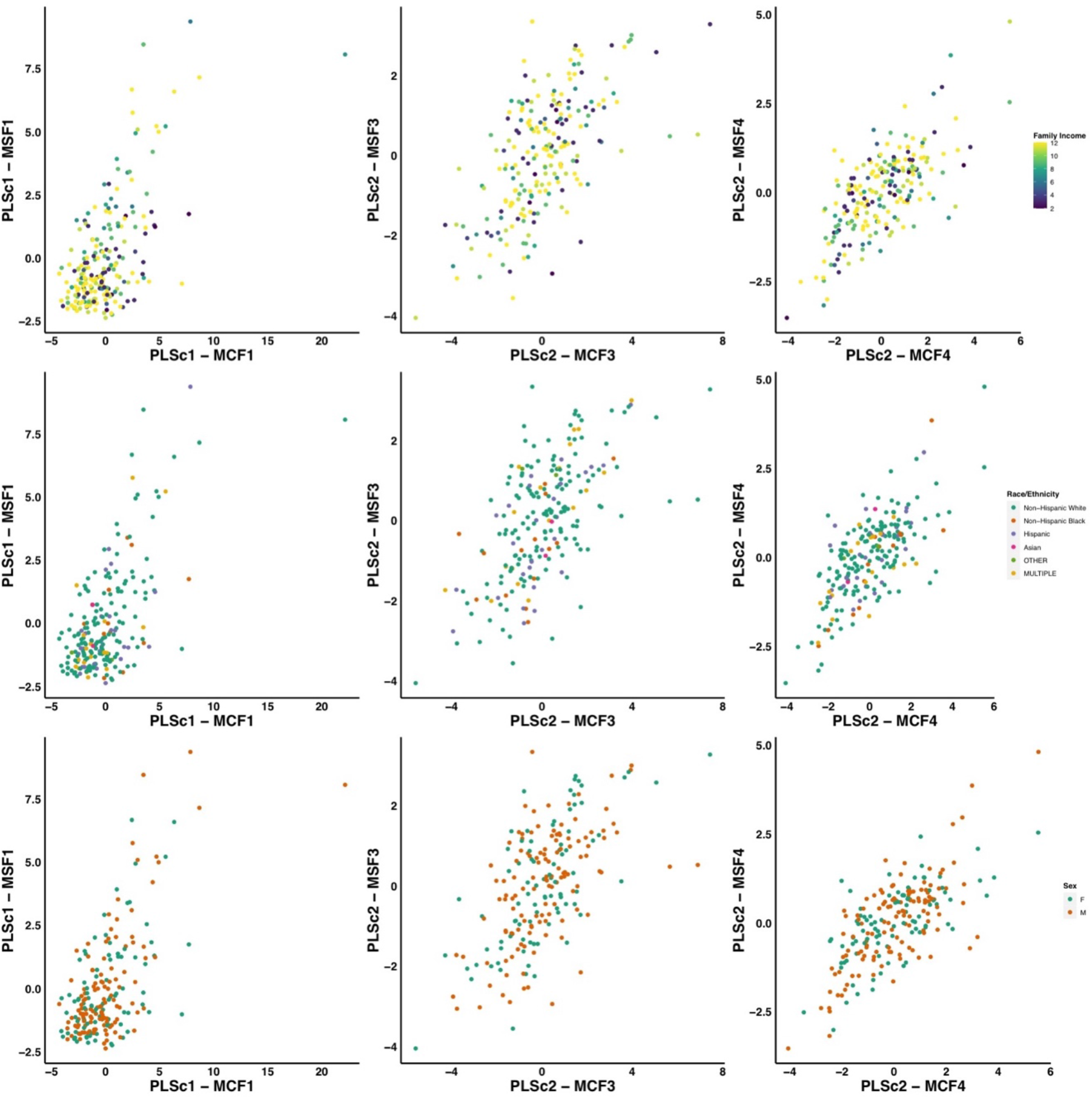
Scatter plots illustrating the expression of three multi-tract multi-symptom pairs (first column: pair 1 from the microstructural complexity PLSc, second column: pair 3 from the axonal density PLSc, third column: pair 4 from the axonal density PLSc), color-coded by total family income (first row), race/ethnicity (second row), and sex (third row).

**Figure S2.**
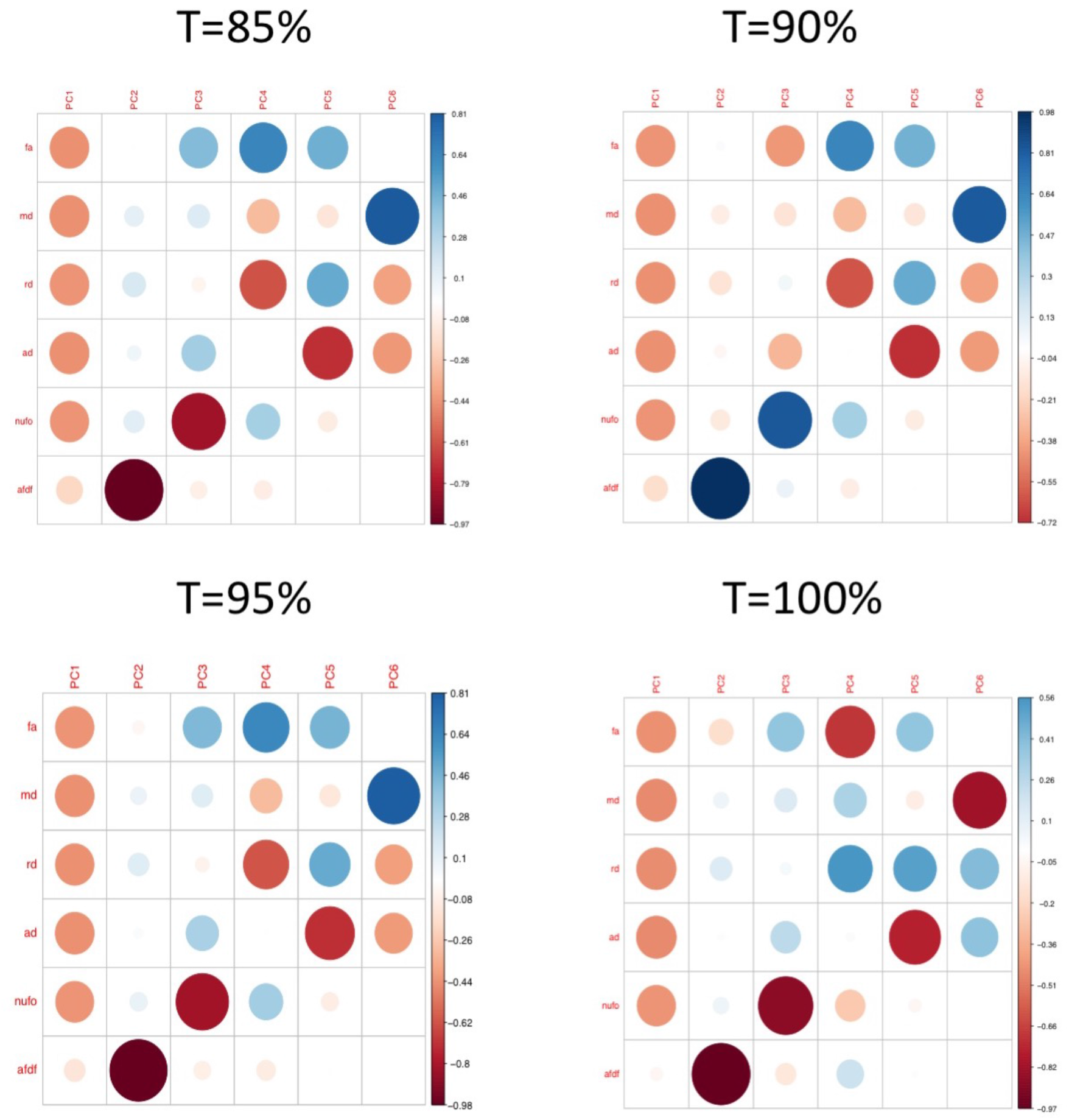
Plots illustrating the weights of each diffusion measure for each principal component for different connectome thresholds (85%, 90%, 95%, 100%). The interpretation of the first two principal components are consistent across thresholds.

**Figure S3.**
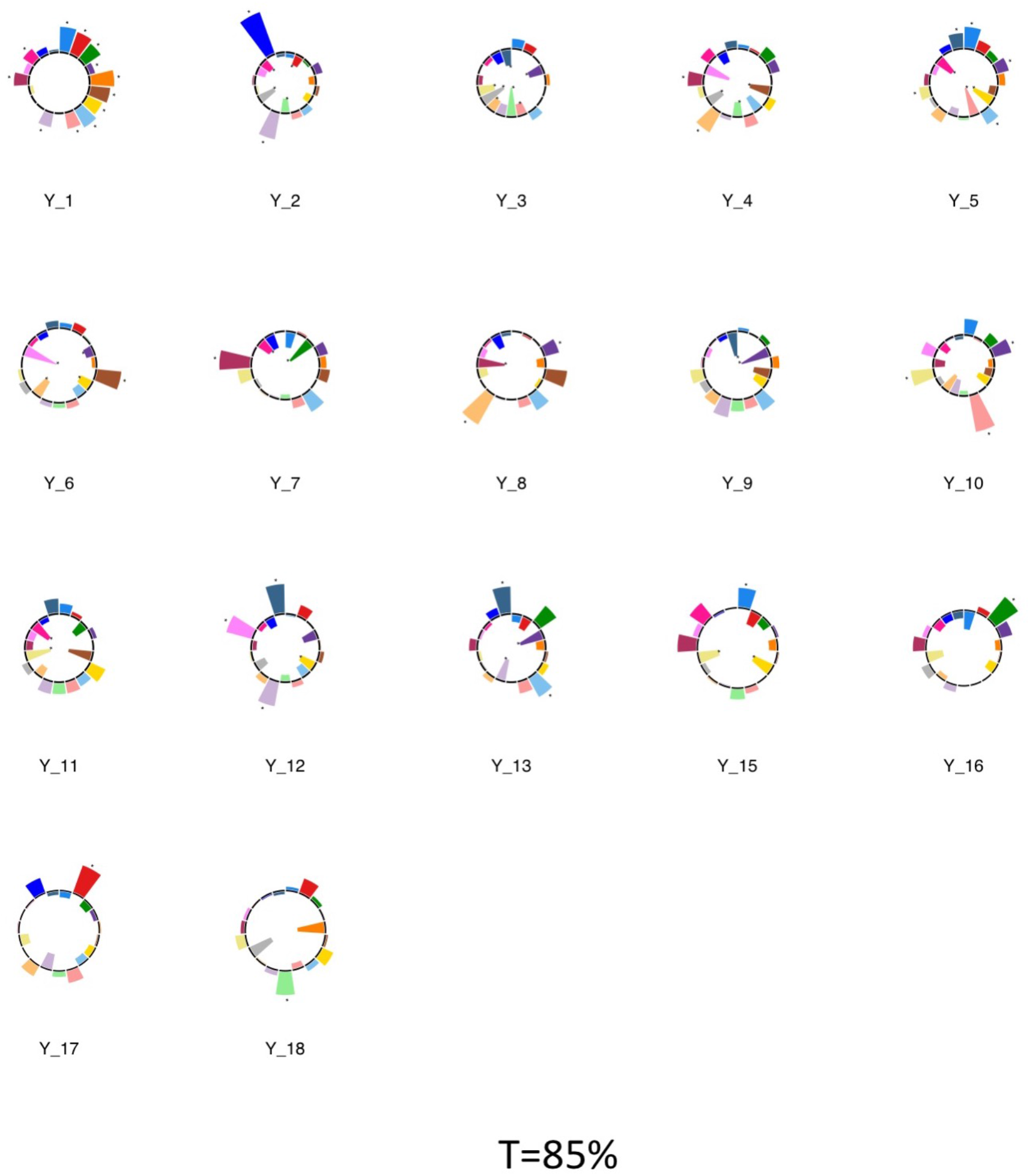
Polar plots illustrating the weights of each symptom measure for every retained multi-symptom feature obtained from the microstructural complexity PLSc performed using all 19 symptom measures as well as connectivity features selected from connectomes thresholded at T=85%. Black stars indicate symptoms that significantly contributed to the multi-tract multi-symptom pair based on bootstrapping analyses.

**Figure S4.**
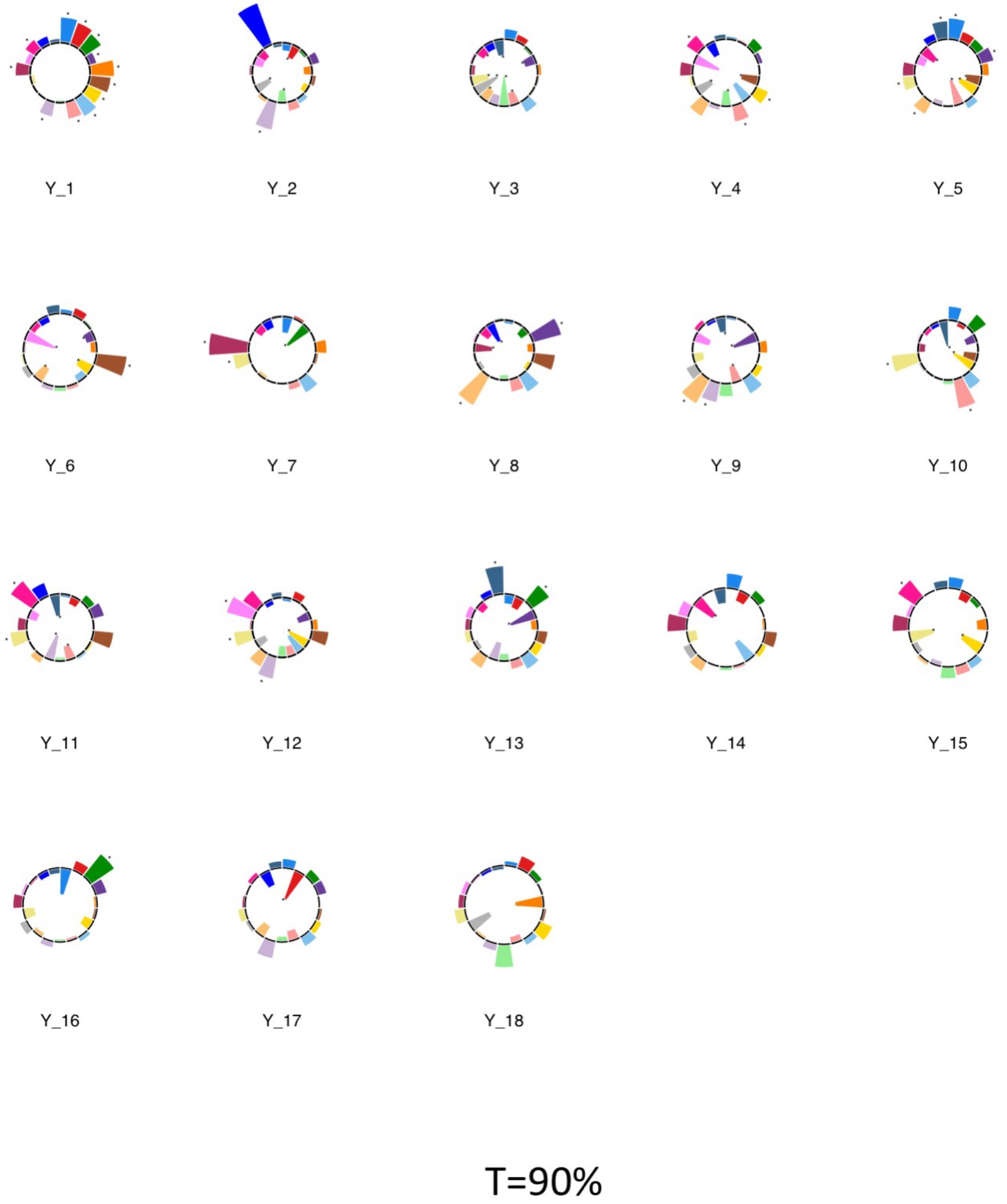
Polar plots illustrating the weights of each symptom measure for every retained multi-symptom feature obtained from the microstructural complexity PLSc performed using all 19 symptom measures as well as connectivity features selected from connectomes thresholded at T=90%. Black stars indicate symptoms that significantly contributed to the multi-tract multi-symptom pair based on bootstrapping analyses.

**Figure S5.**
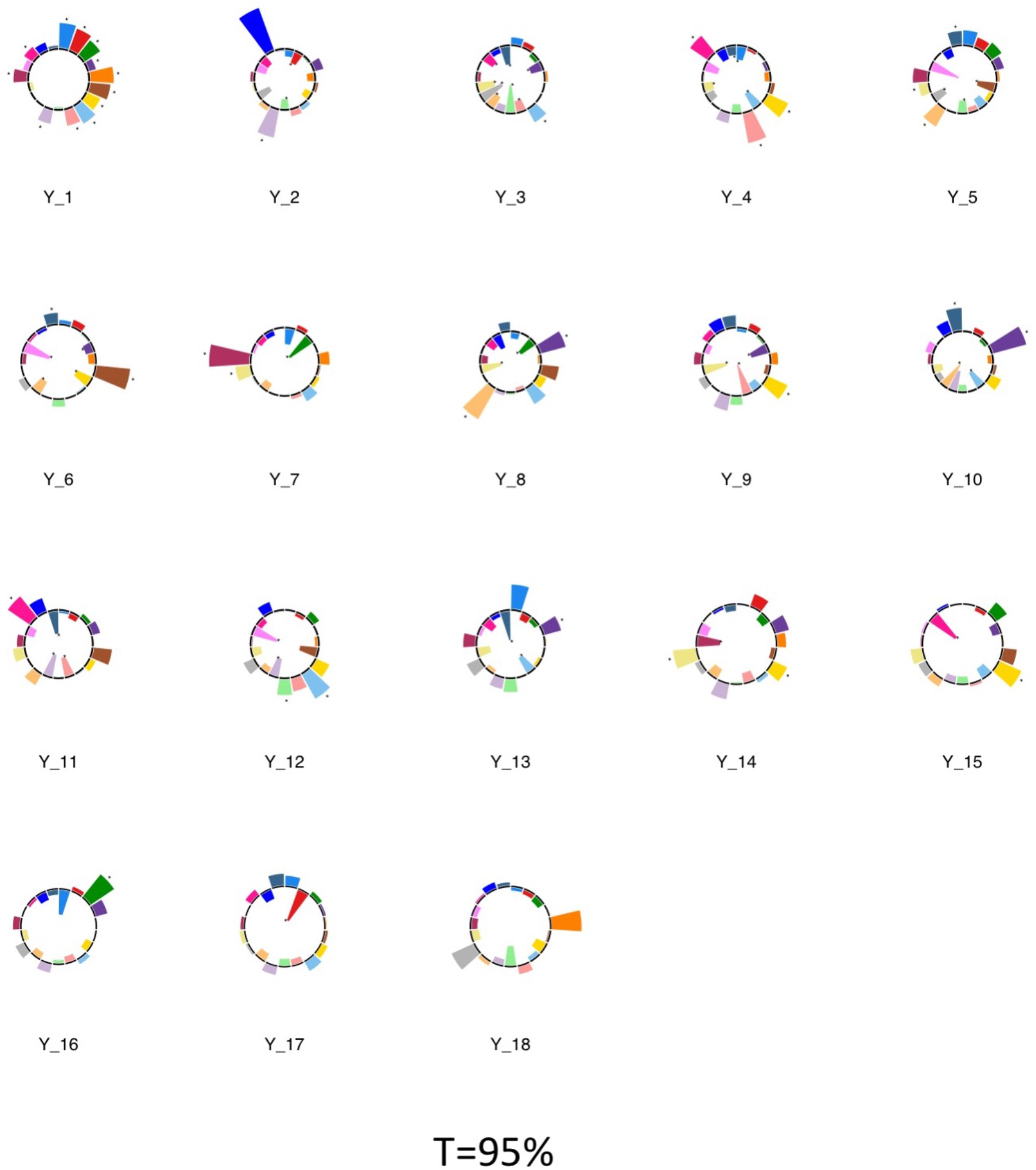
Polar plots illustrating the weights of each symptom measure for every retained multi-symptom feature obtained from the microstructural complexity PLSc performed using all 19 symptom measures as well as connectivity features selected from connectomes thresholded at T=95%. Black stars indicate symptoms that significantly contributed to the multi-tract multi-symptom pair based on bootstrapping analyses.

**Figure S6.**
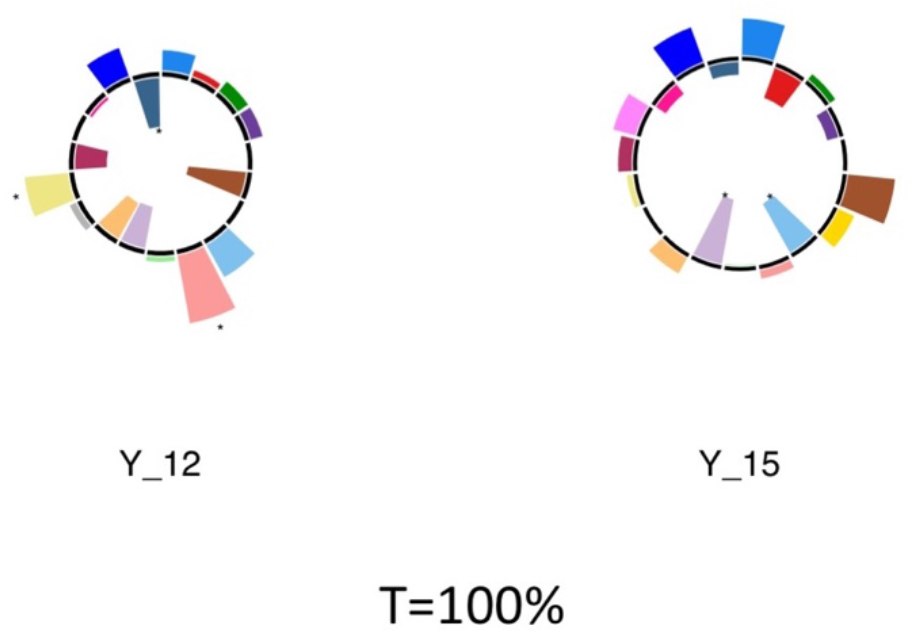
Polar plots illustrating the weights of each symptom measure for every retained multi-symptom feature obtained from the microstructural complexity PLSc performed using all 19 symptom measures as well as connectivity features selected from connectomes thresholded at T=100%. Black stars indicate symptoms that significantly contributed to the multi-tract multi-symptom pair based on bootstrapping analyses.

**Figure S7.**
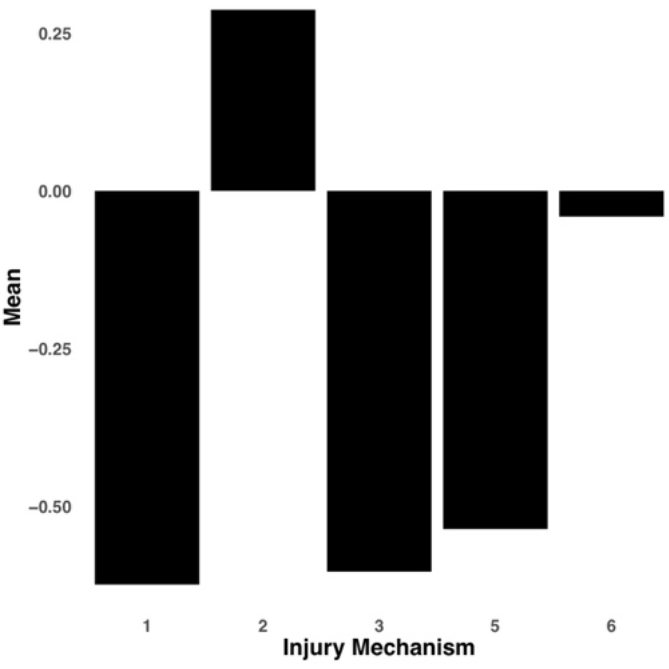
Bar graph illustrating the expression of multi-tract connectivity feature 7 from the microstructural complexity PLSc, averaged according to subgroups of participants defined by Injury Mechanism. 1: Fall/hit by object; 2: Fight/shaken; 3: Motor vehicle collision; 4: Multiple; 5: Unknown.

**Figure S8.**
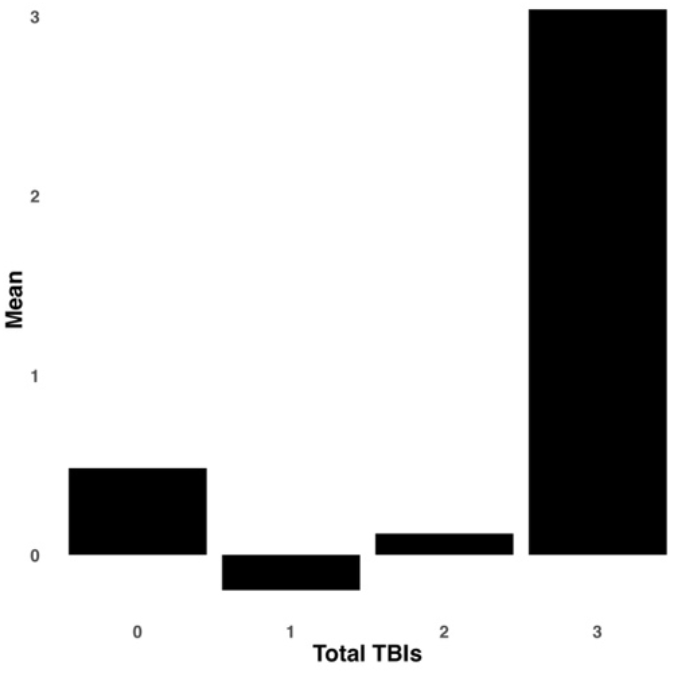
Bar graph illustrating the expression of multi-tract connectivity feature 17 from the microstructural complexity PLSc, averaged according to subgroups of participants defined by Total TBIs. 0: Unknown. Other numbers represent the total number of TBIs.

**Figure S9.**
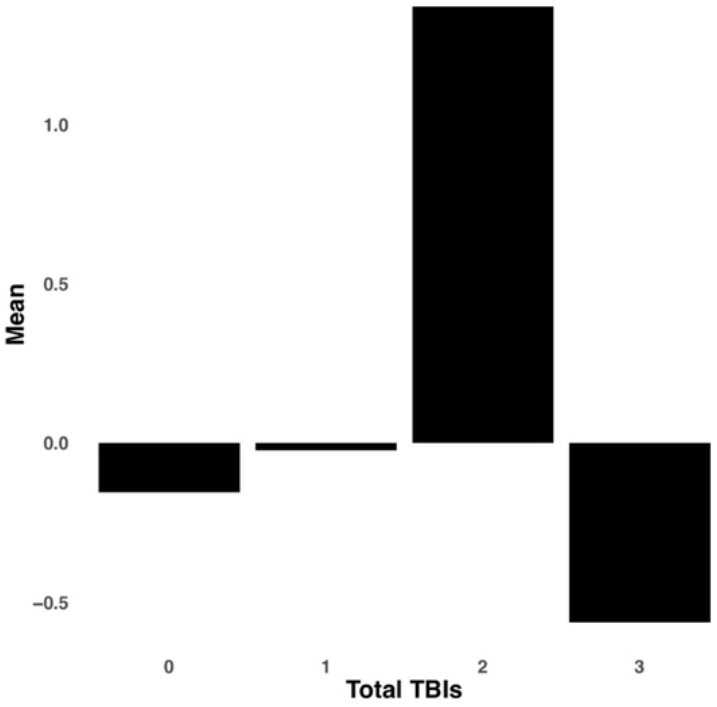
Bar graph illustrating the expression of multi-tract connectivity feature 8 from the axonal density PLSc, averaged according to subgroups of participants defined by Total TBIs. 0: Unknown. Other numbers represent the total number of TBIs.

**Figure S10.**
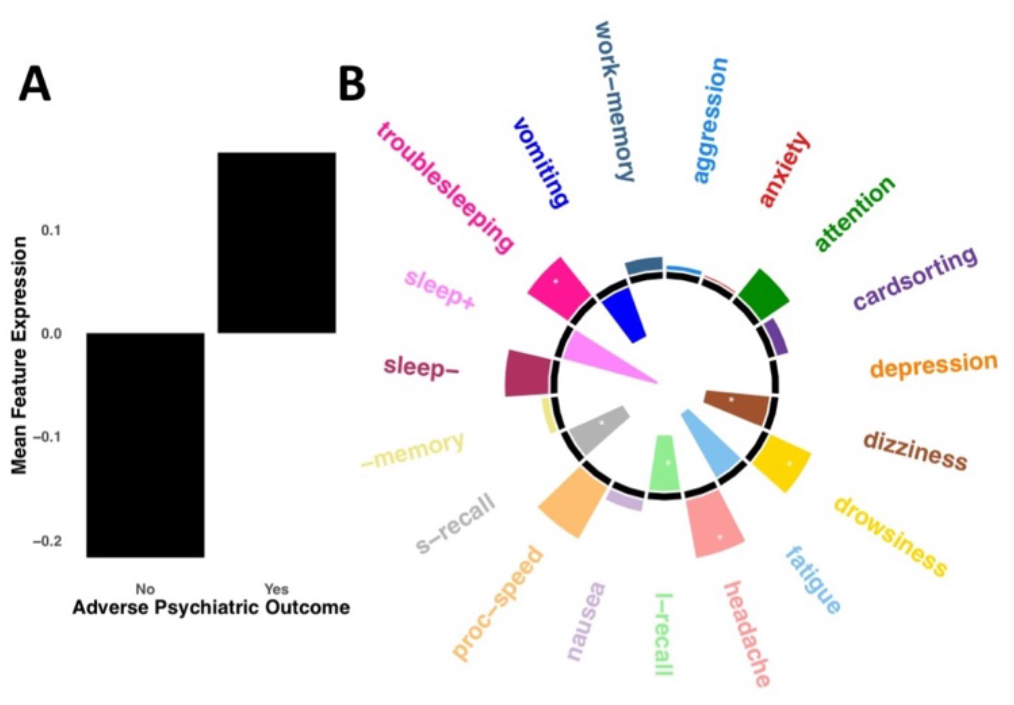
A. Bar graph illustrating the expression of multi-tract connectivity feature 4 from the microstructural complexity PLSc, averaged according to subgroups of participants defined by adverse psychiatric outcome (defined as a current psychiatric diagnosis). B. Polar plot displaying the weights of all 19 symptom-oriented measures for multi-symptom feature 4. Bars pointing away from the circle center illustrate positive weights, bars pointing towards the circle center represent negative weights. White stars illustrate symptoms that were found to significantly contribute to this multi-tract multi-symptom pair based on bootstrapping analyses.

**Figure S11.**
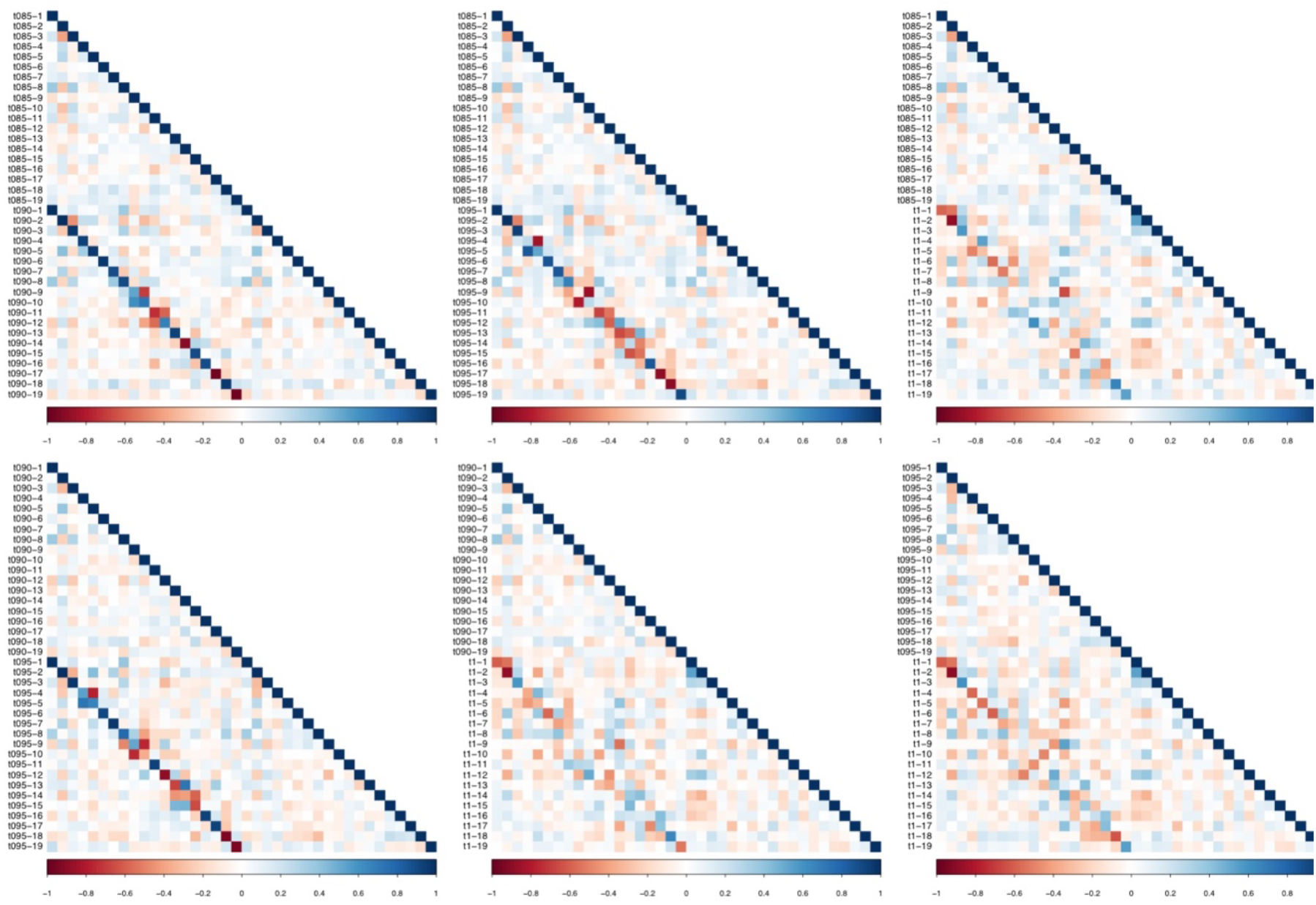
Matrices illustrating correlation coefficients between the expression of every pair of multi-tract connectivity features obtained from the microstructural complexity PLSc. Each matrix illustrates the correlation between features obtained from two PLSc analyses. Each analysis was performed on connectivity features obtained from different connectome thresholds. From left to right, top row: thresholds 85% and 90%, thresholds 85% and 95%, thresholds 85% and 100%. From left to right, bottom row: thresholds 90% and 95%, thresholds 90% and 100%, thresholds 95% and 100%. Given that these matrices are symmetrical, only the bottom triangular is shown. The main diagonals illustrate autocorrelations. These matrices illustrate how corresponding multi-tract connectivity features between thresholds (e.g.: multi-tract connectivity feature 1 from T=85%, multi-tract connectivity feature 1 from T=90%) are highly correlated across most thresholds, except with T=100%, which appears to be most dissimilar from the other thresholds.

**Table S1.** Table listing *p*-values of correlations between the expression of all retained multi-tract connectivity features and the time since the latest injury.

| **Multi-tract**<br>**connectivity**<br>**feature** | **PLSc1** | **PLSc2** |
| --- | --- | --- |
| MCF1 | 0.801 | 0.842 |
| MCF2 | 0.969 | 0.605 |
| MCF3 | 0.202 | 0.0101 |
| MCF4 | 0.415 | 0.87 |
| MCF5 | 0.193 | 0.529 |
| MCF6 | 0.406 | 0.718 |
| MCF7 | 0.601 | 0.709 |
| MCF8 | 0.711 | 0.3 |
| MCF9 | 0.222 | 0.399 |
| MCF10 | 0.0495 | 0.554 |
| MCF11 | 0.521 | 0.538 |
| MCF12 | 0.487 | 0.968 |
| MCF13 | 0.19 |  |
| MCF14 | 0.886 | 0.187 |
| MCF15 | 0.07 |  |
| MCF16 | 0.688 | 0.415 |
| MCF17 | 0.0582 |  |
| MCF18 | 0.638 |  |

PLSc1: Microstructure complexity PLSc; PLSc2: Axonal Density PLSc.

